# Considerations regarding a diagnosis of Alzheimer’s Disease before dementia: a systematic review

**DOI:** 10.1101/2021.09.16.21263690

**Authors:** Jetske van der Schaar, Leonie N.C. Visser, Femke H. Bouwman, Philip Scheltens, Annelien L. Bredenoord, Wiesje M. van der Flier

**Author notes:** corresponding author: Jetske van der Schaar, Alzheimer Center Amsterdam, Department of Neurology, VU University Medical Center, Amsterdam UMC, De Boelelaan 1118, 1081 HZ Amsterdam, The Netherlands.

## Abstract

**Introduction:** The NIA-AA research framework proposes a purely biological definition of Alzheimer’s Disease (AD). This implies AD can be diagnosed based on biomarker abnormalities. While this brings opportunities, it also raises challenges.

**Methods:** We conducted a systematic review by searching PubMed for publications on conveying AD biomarker results to individuals without dementia. Content was analyzed inductively.

**Results:** We included 25 publications. From these we extracted 26 considerations, which we grouped according to their primary relevance to a clinical, personal, or societal context. Clinical considerations include (lack of) validity, utility, and disclosure protocols. Personal considerations cover psychological and behavioral implications, as well as the right to (not) know. Societal considerations comprise the risk of misconception, stigmatization, and discrimination. Overall, views were heterogeneous and often contradictory.

**Discussion:** Perceptions on a diagnosis of AD before dementia vary widely. Empirical research is required, taking perspectives of medical professionals and the general public into account.

## 1 INTRODUCTION

The pathophysiological cascade of events in Alzheimer’s disease (AD) starts twenty to thirty years before dementia [1-3]. Nowadays it is possible to detect this pathology *in vivo*, using biomarkers. The National Institute on Aging and Alzheimer’s Association Research Framework operationalized AD as a biological construct characterized by evidence of amyloid plaques, tau tangles and neurodegeneration, irrespective of clinical expression [4]. This implies that AD can be diagnosed before dementia, in individuals with mild cognitive impairment, subjective cognitive decline, or normal cognition.

This development has sparked a heated debate. Should physicians tell individuals without dementia that they have AD? This may lead to distress [5] as a precise prognosis cannot be given and there is no curative therapy (yet). Alternatively, can physicians withhold information on underlying disease from patients, just because they don’t fulfil clinical dementia criteria? While perhaps avoiding anxiety, this would deprive patients of the opportunity to adopt a risk-reducing lifestyle, prepare for the future, or participate in dementia-prevention trials [6].

The numbers of persons living with preclinical AD are large [7] and many wish to learn their biomarker results [8]. A first estimation suggests the prevalence of AD may be three times higher when based on a biological rather than a clinical definition of the disease, illustrating the potential magnitude of consequences [9]. With the conditional approval of a first disease-modifying therapy [10], this discussion is more relevant than ever.

We aim to provide an overview of considerations regarding the disclosure of AD pathology before dementia.

## 2 METHODS

We conducted a systematic literature search following PRISMA guidelines [11]. Our broad query combined variations on the terms “Alzheimer’s” AND “disclos*” OR “diagnos*” AND “predementia” AND “biomarkers”, using controlled standardized keywords (MeSH) as well as (truncated) free text terms. We searched PubMed for results published before 10 December 2020 and scanned reference lists of identified articles.

All articles in English presenting theoretical data, i.e. ethical concerns, psychosocial consequences and societal implications of disclosing amyloid and/or tau results to individuals in AT(N) stages 1-3 [4] were eligible, except editorials. Publications on later stages and other types of dementia or neurodegenerative diseases were excluded, as well as those primarily focused on trial design or genetic risk.

Two authors independently screened all titles and abstracts. The remaining articles were assessed for eligibility based on full text. In case of discrepancy, consensus was reached after discussion. We performed inductive content analysis, [12] using MAXQDA-software, to extract considerations and underlying arguments. Based on the data, these were further subcategorized in meaningful clusters. An additional classification was made according to the key ethical principles of medical ethics, i.e., beneficence, non-maleficence, justice and autonomy [13].

## 3 RESULTS

Our initial search yielded 3985 records. After applying selection criteria (Figure **1**. Flow diagram of study selection), we included 25 articles.

**Figure 1.**
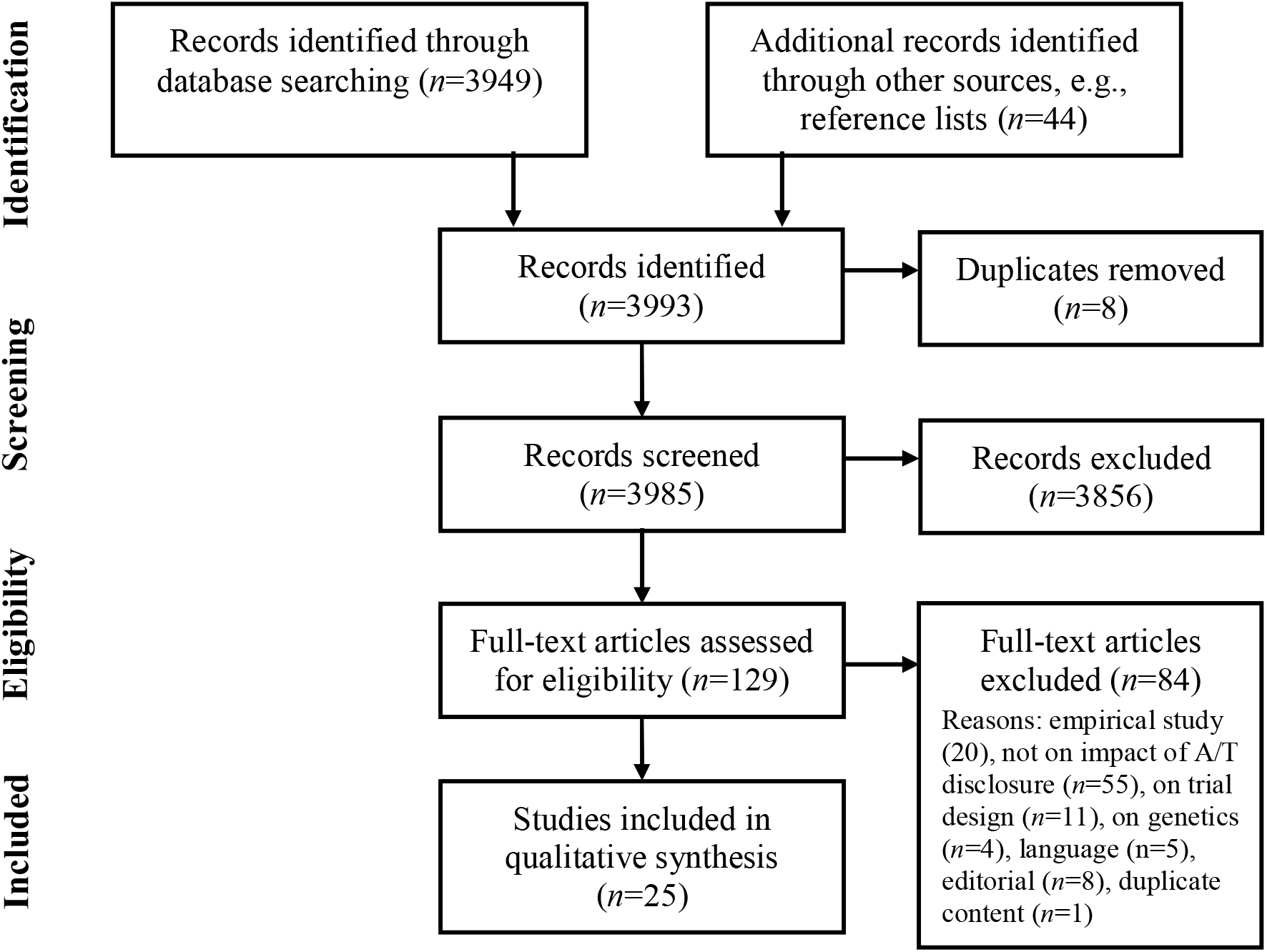
Flow diagram of study selection

**Figure 2.**
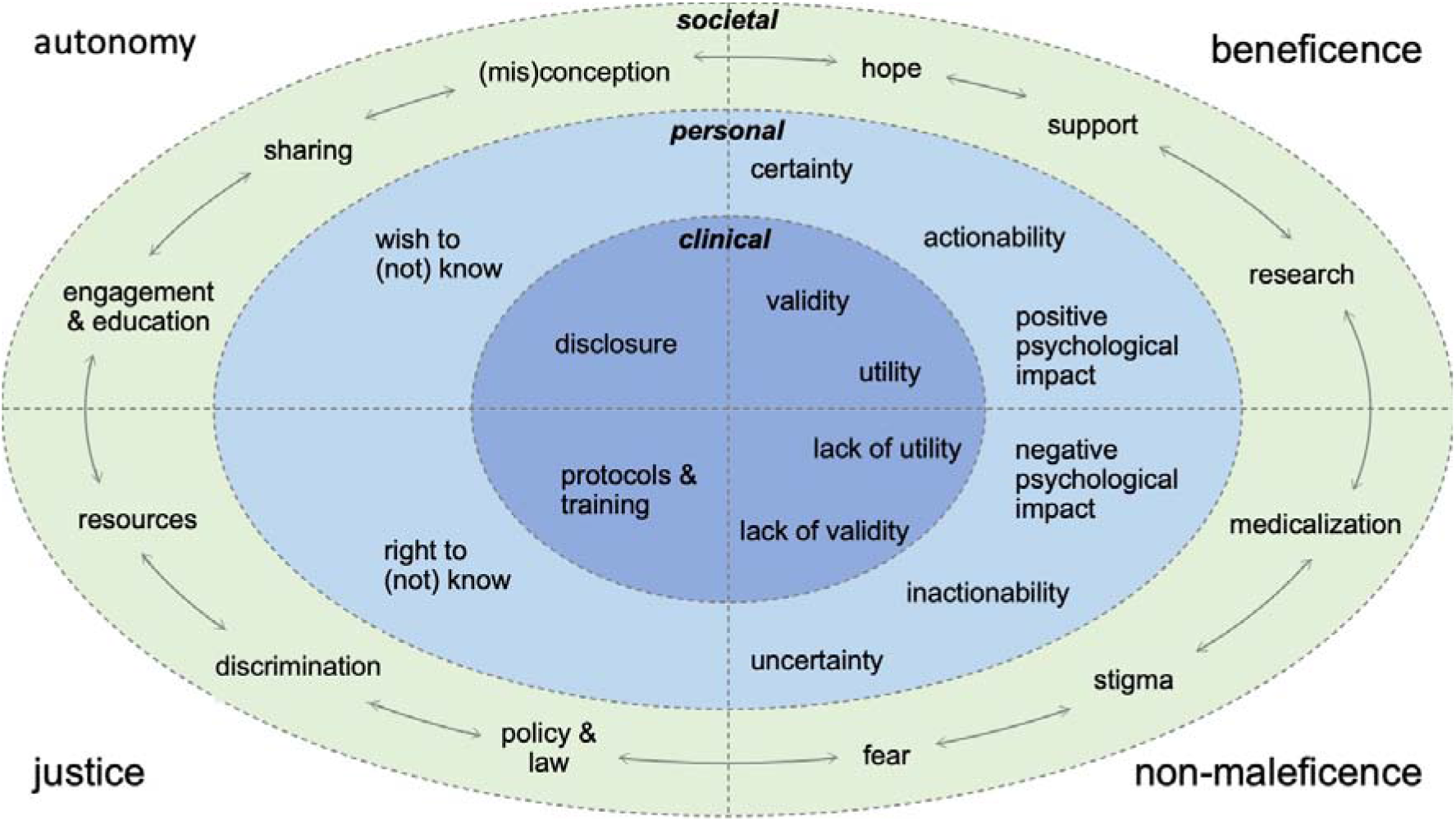
Visual overview of considerations Visual overview of 26 considerations extracted from the included literature, categorized based on the clinical, personal, or societal context they relate to and the four basic principles of biomedical ethics: beneficence (doing good), non-maleficence (avoiding harm), justice (ensuring fair distribution of resources in accordance with the law) and autonomy (allowing free, informed and deliberate decisions). Contested issues, e.g., (in)actionability, are ranked under beneficence as well as non-maleficence to reflect the theoretical debate and their subjective nature. Arrows illustrate the interaction between societal considerations, e.g., that sharing test results can lead to support but also stigma and discrimination. The visual overview highlights the tension between clinicians’ responsibility to weigh the benefits and risks and prevent unnecessary suffering, versus individuals’ right to self-determination.

From these, 26 unique considerations were extracted (Table 1), and further categorized according to their primary relevance to a clinical, personal or societal context.

**Table 1.**
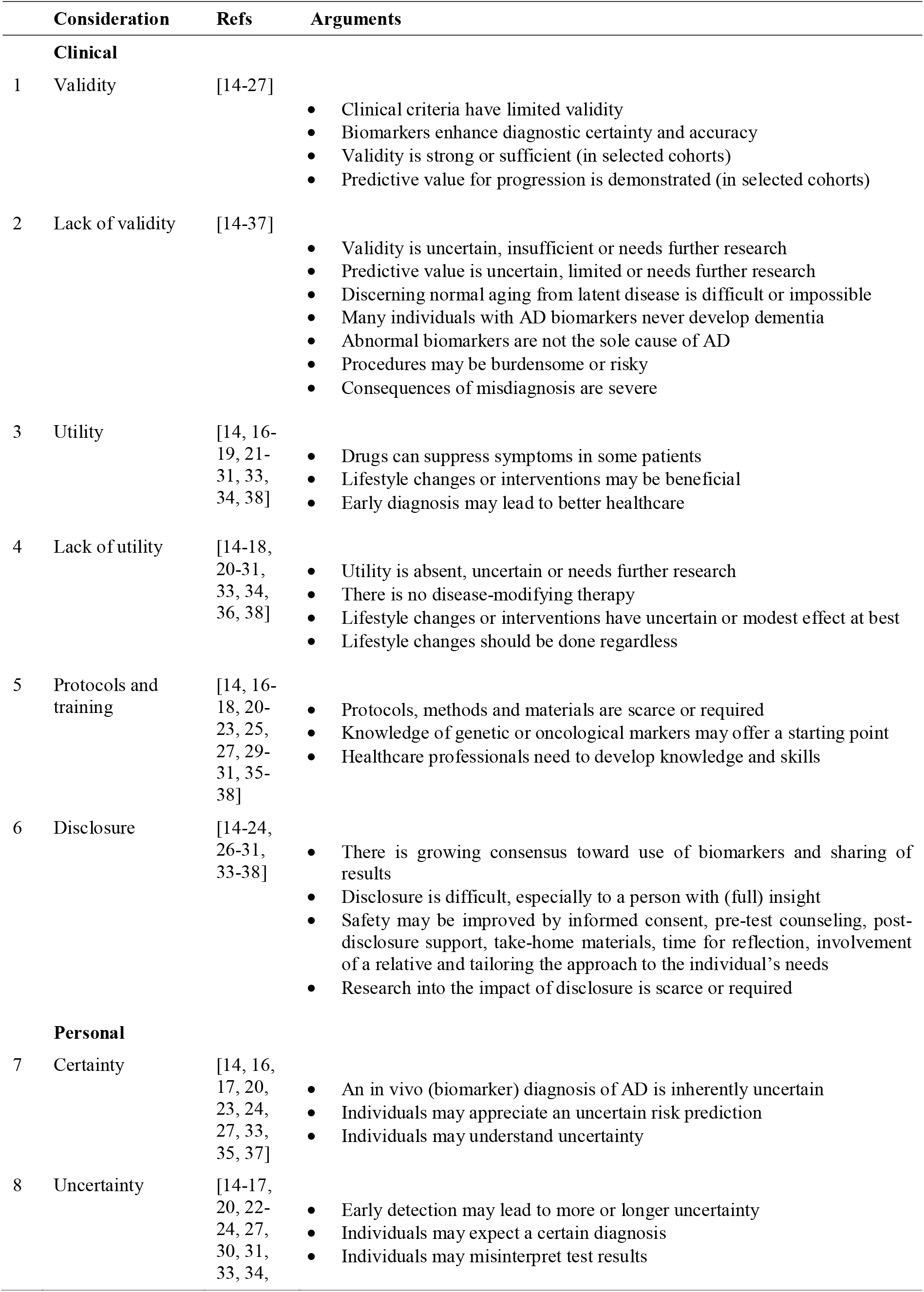
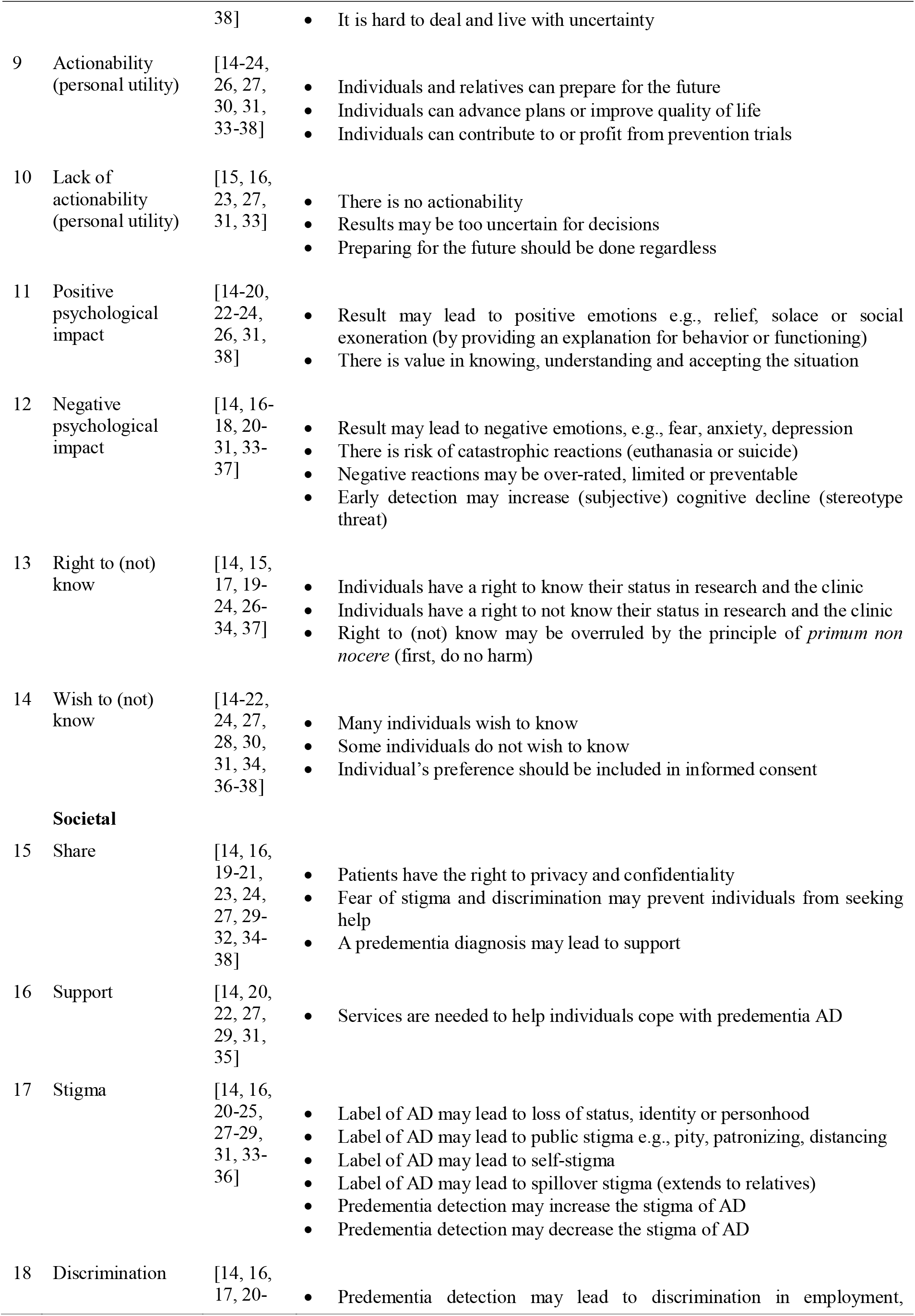
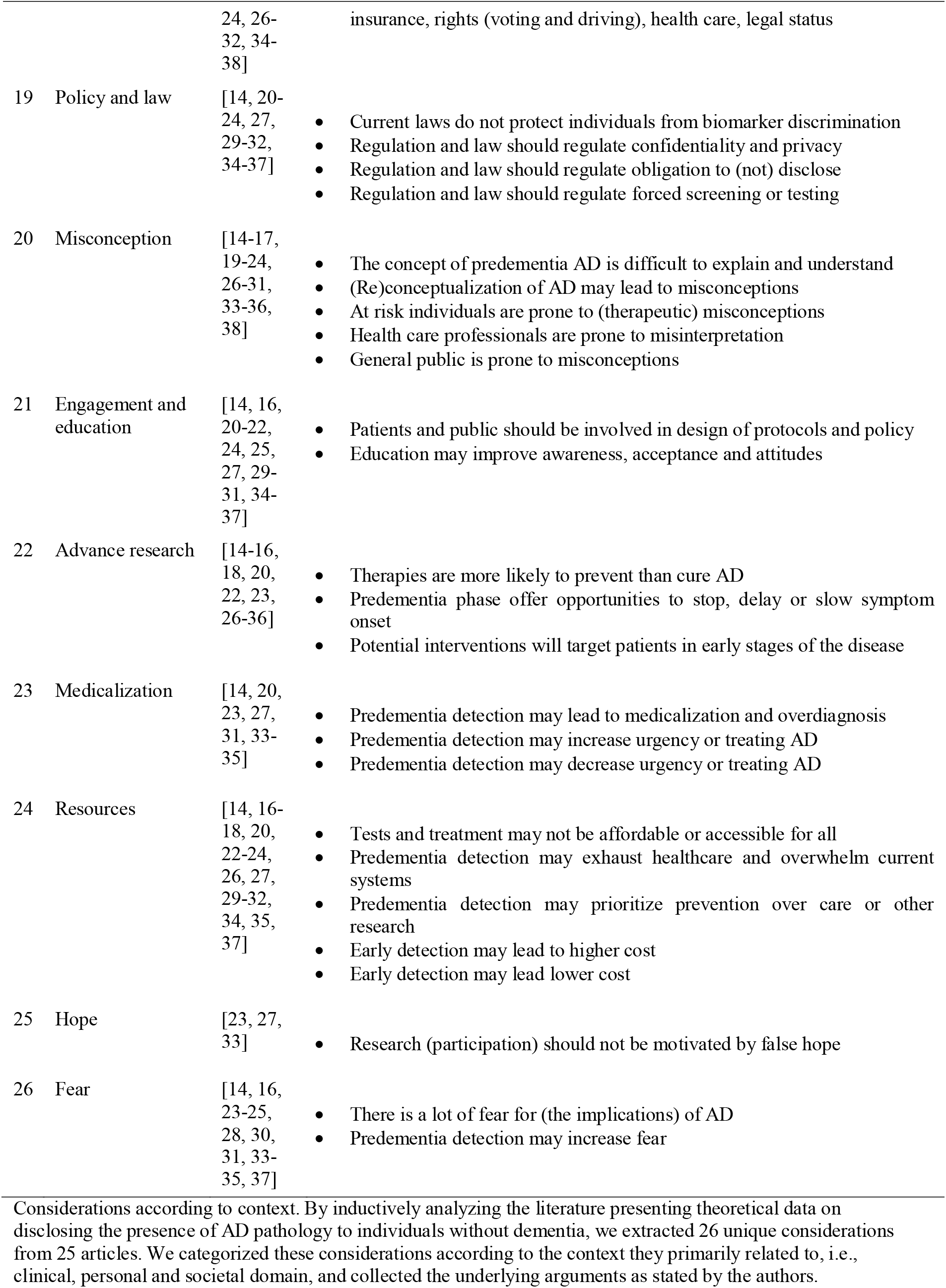
Clinical, personal and societal considerations

### 3.1 Clinical considerations

#### 3.1.1 (Lack of) clinical validity

Although most authors acknowledged that biomarker information enhances diagnostic accuracy [14-24] and validity is strong in selected cohorts [17-21, 25, 26], the predictive value was debated [14-34]. Many discussed the difficulty to discern normal aging from latent AD [14-16, 18, 20, 22, 25, 26, 28-30, 32, 33, 35], as the majority of cognitively healthy elderly with abnormal biomarkers never develop dementia [14-16, 20-23, 25, 26, 28-30, 32, 33, 36], since AD is multi-factorial [16, 18, 25, 26, 30, 34]. It was argued that procedures are not without burden or risk [14, 16, 18, 20, 21, 23, 24, 33] and consequences of incorrectly labeling people as ‘patients in waiting’ [24, 33] could be severe [14, 16, 24-27, 30]. Most authors concluded that biomarker criteria require final demonstration of validity in populations without dementia [14-23, 25, 26, 29, 30, 32-34, 37].

#### 3.1.2 (Lack of) clinical utility

Clinical utility was contested, mainly due to the absence of a disease-modifying therapy [14-16, 18, 20, 21, 23, 24, 26, 27, 30, 33, 34, 36, 38], and limited effectiveness of symptom suppressing medications, which has not been demonstrated in predementia stages of AD [21, 24]. On the other hand, several authors suggested lifestyle interventions could delay cognitive impairment [14, 16, 17, 21-31, 33], although evidence on their effectiveness remains inconclusive [14, 15, 21, 30], and according to some, such health improvements should be pursued regardless of one’s biomarker status [16, 23, 30]. Lastly, it was argued that early detection can improve patient care, e.g., by offering an explanation for concerns, anticipating medical needs, and facilitating access to support [14, 16, 18, 19, 22-24, 26, 27, 31, 34]. Nonetheless, others did not consider biomarker information medically meaningful [15, 16, 20-25, 28-31].

#### 3.1.3 Protocols and training

Many papers addressed the need for guidance regarding who to test, what findings signify, whether to disclose and how [17, 18, 20-22, 27, 30, 35-37]. Knowledge from the fields of oncology and genetic testing may offer a good starting point [14, 20, 23, 26, 27, 29, 35-37]. The literature referred to development of standardized processes and materials for disclosing biomarker levels in prevention trials [17, 29, 31, 35, 36], but reported ‘a complete lack of studies in a clinical setting’ [17]. In addition, it was emphasized that the required skills cannot be assumed, so medical professionals would profit from training in risk communication [14, 16, 18, 20, 22, 27, 35, 36, 38].

#### 3.1.4 Disclosure

Conveying an AD diagnosis was considered daunting, especially to persons with full insight [15, 17, 20-24, 28, 29], as knowledge of the impact is scarce, yet urgently required [14-17,20, 22-24, 26, 29-31, 33, 35, 36]. Recommendations derived from research on genetic risk covered informed consent, pre-test screening, education and counseling, sufficient time for consultation and reflection, take-home materials, involving a relative and tailoring to individuals [14, 15, 17, 19-22, 24, 26, 27, 29-31, 34-38].

### 3.2 Personal considerations

#### 3.2.1 (Un)certainty

While an abnormal biomarker result provides certainty on the presence of brain lesions, it was also argued to cause uncertainty about the eventuality of cognitive decline [14, 20, 23, 24, 31, 33], which was suggested to be hard to deal with [20, 21, 31]. While some authors felt individuals might appreciate and understand such indefinite risk [17, 23, 33, 35, 37], many others feared that it would be misinterpreted as an inevitability [14-16, 20, 22, 24, 27, 30, 34,38].

#### 3.2.2 (In)actionability

Proponents stated that awareness of having AD without dementia allowed individuals and relatives to prepare for the future by arranging private, professional, financial and legal matters, obtaining long-term care insurance, writing advance directives or making end-of-life decisions [14-24, 26, 27, 30, 31, 33, 35-38], participating in prevention trials [14-18, 20, 22-24, 34, 36], or retiring early and enjoying time left [14, 23, 38]. Opponents believed these things should be done anyway [15, 16, 23], and questioned whether the predictive power was sufficient to substantiate far-reaching decisions [15, 27, 31].

#### 3.2.3 Negative / positive psychological impact

Several publications observed the absence and even presence of AD lesions might offer relief, solace, or an explanation of symptoms [17-19, 22-24, 26, 31, 38]. Knowing what is going on was thought to be of value in itself [14-17, 20, 22-24, 26, 31]. Yet, nearly all papers mentioned adverse emotions including fear, anxiety and depression [14, 16-18, 20-31, 33-37], or suicidal ideation [17, 18, 20, 22, 23, 27, 29, 31, 34, 36, 37], despite reason to believe negative reactions may be over-rated, limited or temporary [17, 18, 20, 22, 23, 27, 29, 31, 34, 36, 37]. Another worry was the risk of stereotype threat or nocebo reaction, where the knowledge of susceptibility leads to the associated behavior or a decrease in memory functioning [14, 17, 23, 27, 30, 33, 35].

#### 3.2.4 Right to (not) know

On moral and legal grounds an individual’s request to access one’s personal data must be granted, be it in a research trial or clinical practice [15, 17, 19-24, 26-28, 30, 33, 37]. Likewise, a refusal to be informed of such information must be respected as well [14, 15, 20, 21, 23, 24, 27, 34]. However, it was argued that this fundamental right can be in conflict with physicians’ oath of *primum non nocere* (first, do no harm) and in some cases could or even should be overruled [14, 17, 19-24, 26-32].

#### 3.2.5 Wish to (not) know

Authors reported that many individuals express a desire to receive risk information [15-17, 22, 24, 27, 28, 30, 31, 37], while some might prefer to remain ignorant of uncertain odds [16, 17, 21, 24, 28, 31]. It was recommended that extensive and ‘truly’ informed consent [30] should record a patient’s preference and list which other persons and authorities will be notified [14, 15, 18, 20, 21, 24, 27, 30, 31, 34, 36-38].

### 3.3 Societal domain

#### 3.3.1 Sharing

As individuals have the right to privacy and confidentiality [14, 19-21, 23, 24, 27, 29-32, 34-38], it was emphasized that diagnostic information should not be released to relatives [24, 34] or third parties [14, 20, 21, 23, 24, 29-32, 34, 35, 37] without their consent and against their interest, although in case of driving, physicians may be obliged to report this to relevant authorities [36]. It was also mentioned that fear of stigma could prevent patients from voicing their concerns and seeking help, while doing so may also yield support [14, 16, 20, 23, 27, 31, 34, 35].

#### 3.3.2 Support services

Apart from pre- and post-diagnostic counseling [14, 17, 20-22, 24, 27, 29-31, 34-38], literature addressed the need of support services for people along the continuum of AD, including assistance in personal, social and healthcare needs, and monitoring of professional, financial and legal capacities [14, 20, 22, 25, 27, 29, 31, 35, 37].

#### 3.3.3 Stigma

According to the literature, public stigma ranges from patronizing attitudes to social distancing, exclusion, and isolation [14, 23, 25, 27-29, 31, 33, 35, 36]. This induces self-stigma, when pejorative views are internalized as feelings of shame, lowered self-esteem, and inferiority [14, 23, 29, 31, 33, 35]. In addition, spillover-stigma is detrimental to family members [14, 27, 28, 31, 35]. An increase of predementia patients was expected to expand stigma [14, 23, 29, 33, 35, 36], conversely, normalization was assumed to dilute it as well [14, 20, 35].

#### 3.3.4 Discrimination

Individuals with an AD diagnosis and risk of dementia were considered vulnerable to discrimination, affecting their professional position, insurance fees, legal status, civil rights (driving and voting) and financial capacity [14, 16, 17, 20-24, 26-32, 34-38].

#### 3.3.5 Policy and law

Current legislation, such as the United States Genetic Information Non-Discrimination Act and the Americans with Disabilities Act, does not adequately protect individuals with predementia AD [20, 22, 27, 32, 35, 36]. Several authors advocated for regulation of confidentiality and privacy, preclinical screening, and obligatory disclosure for persons with high responsibility [14, 20, 21, 23, 24, 27, 29-32, 34-37].

#### 3.3.6 Misconception

Authors observed that a symptomless condition with uncertain prognosis is hard to grasp [14, 17, 20, 22-24, 27, 28, 34], and the changing meaning of ‘AD’ could lead to incorrect interpretations [14, 21, 24, 28, 29, 31, 33], especially since at-risk individuals are prone to misconceptions [14-17, 22, 23, 27, 30, 33, 34, 38], healthcare providers apply different interpretations of disease criteria [14, 16, 17, 19, 22, 26-28, 30], and the general public is influenced by dementia myths and the media’s portrayal of AD patients as ‘dehumanized shells’ [14, 29, 35].

#### 3.3.7 Education and engagement

Many publications stress that individuals’ perceptions can change after intervention or experience [15, 17, 27, 30, 33, 36], and public dialogue may improve awareness and attitudes [14, 22, 24, 25, 27, 29, 30, 34, 35, 37]. Moreover, patients of all cultures should be involved in development of protocols and policy, to represent their own views, improve research and decrease stigma [14, 16, 20-22, 24, 27, 29, 31, 34, 35].

#### 3.3.8 Resources, opportunities, and costs

Authors worried that predementia testing may not be accessible and affordable for all [14, 16, 23, 24, 26, 29, 35], individuals with minimal symptoms could strain healthcare services [14, 22, 23, 35] and a focus on prevention research might come at the expense of patients with advanced AD [14, 16, 20, 34, 35]. Thus, the emotional and financial burden could rise substantially [14, 16-18, 20, 23, 24, 29-32, 34, 35], or drop considerably when patients live longer at home, at-risk participants lower trial costs and medication becomes available [14, 16, 18, 20, 23, 26, 27, 31, 37].

#### 3.3.9 Medicalization

Expanding the criteria for AD raised concerns of tipping the scales from under- to overdiagnosis [14, 20, 23, 27, 31, 34, 35]. Paradoxically, normalization was reasoned to result in marginalization, but also argued to increase the urgency to develop disease-modifying therapies [14].

#### 3.3.10 s Advance research

The primary consideration behind the new criteria is to prevent individuals with AD from developing dementia, as early interventions are hypothesized to have better chances of success. Finally, authors were wary of inflating unsubstantiated hope [23, 27, 33] and/or further fueling already widespread fear [14, 16, 23-25, 28, 30, 31, 33-35, 37]. Both were primarily regarded as vulnerabilities and impediments to rational decision making [23, 33, 34].

### 3.4 Key principles

In substantiating their arguments, many authors invoked the key principles of medical ethics: beneficence, non-maleficence, justice, and autonomy [14, 15, 17, 19-31, 34-38]. **Error! Reference source not found**. visualizes all 26 considerations organized by the context they relate to and the principle that was most applicable.

## 5 DISCUSSION

We found 26 considerations relevant to disclosing a diagnosis of AD to individuals without dementia. These concerns, constraints and implications relate to clinical, personal, and societal contexts. Many constitute direct opposites, such as certainty versus uncertainty, reflecting the heated debate among stakeholders. This duality is concordant with findings from a recent study on patients’ views regarding early AD diagnosis, reporting not only great variety between individuals but also profound ambivalence within individuals [39]. For example, while a diagnosis can provide certainty on what is going on, it can also bring uncertainty on what to expect. Thus, it may not be one or the other, as both sides can be true to some extent, and perspectives may change over time. This illustrates the ardent need of empirical evidence and clinical recommendations on a biomarker-diagnosis of AD. Since market access has been granted to a first disease-modifying therapy [10], the urgency is even greater, as practitioners, patients and society are presented with novel opportunities and challenges.

Particularly with respect to clinical validity, comparison of statements was hampered by a disparity of definitions. In the absence of a gold standard, authors evaluated the accuracy of the biomarker framework [40] according to various views on the true state of AD. Findings were based on different criteria of clinical symptoms, pathological findings, and/or biological changes. While all models have value, they are not interchangeable. Moreover, studies suggest the scientific dissensus on nosology and the shifting meaning of AD create confusion [41-44]. This emphasizes the need for a common concept and language of AD [45]. An underlying and fundamentally contested conundrum is whether individuals with normal cognition, but abnormal biomarkers are ill. Based on research criteria they have AD, but judging by clinical standards they are not sick [21, 26, 29-31, 36, 46]. Two of the included papers evaluate the conceptual validity according to theories of health and disease [25, 33]. The authors reasoned that the signature of amyloid and tau does not represent a singular disease, nor a statistical deviation from normal ageing in the elderly. They concluded that people without symptoms should not be diagnosed as ‘patients-in-waiting’, but considered as persons at-risk [33]. Yet this is not about screening unsuspecting populations. Neither is the phase before dementia entirely without symptoms; individuals present at memory clinics because they experience symptoms in their daily lives, albeit subtle or mild [47, 48]. They wish to learn what’s wrong and they have a right to know. In the field of oncology, it is common to diagnose patients with cancer (in situ), regardless of signs or complaints. The same goes for conditions like hypertension and diabetes mellitus. An apparent difference is the lack of disease-modifying interventions for AD. Some ethicists apply a ‘pragmatic view’ to AD, stating that without preventive medication early detecting may do more harm than good [33]. This view might change with the recent conditional approval of a first disease modifying treatment by the FDA.

Overall, literature tended to concentrate on putative adverse implications. Notably, repeatedly mentioned worries about conforming to stereotypes or nocebo reactions were substantiated by evidence from a single study on disclosure of genetic risk [49]. Although inherited susceptibility is beyond the scope of our review, extensive research on the impact of revealing an increased probability [50-57] or absolute certainty [58-60] of developing dementia has demonstrated that catastrophic outcomes are rare, knowledge of the test results does not affect cognition and participants also perceive benefits. So far, evidence on disclosing biomarkers information is limited, but the few available studies suggest it is safe and actionable [61-66]. However, stigmatization and discrimination are concerns that need further scrutiny [67, 68]. More importantly, it should be noted these findings are based on a selection of individuals, willing to participate and learn their disposition to develop dementia. There is a lack of racial, ethnical, cultural, social, economic, and environmental diversity in study populations. [69]. More empirical research is required, evaluating both harms and benefits, taking perspectives of individuals from all groups into account. The key principles of medical ethics, i.e. beneficence, non-maleficence, justice and autonomy, were frequently invoked to decide whether a predementia diagnosis of AD is justified. However, applying the ‘four principle approach’ may unduly simplify a complicated matter [70]. The framework relies on the notion of a common morality, while the interest, motivation, and implications of biomarker testing are inherently deeply personal [71]. Yet patients’ perspectives and circumstances are under-represented in the theoretical discourse. Rather than risking paternalism by imposing the moral right to know or not know on all, we need a tailored approach to respect the values of each individual. Future research should illuminate which personal factors influence people’s preferences for medical information, as well as the psychological and social implications of disclosing test results. It is pivotal to engage and educate all stakeholders to enable informed (and shared) decision making and empower individuals in choosing what is best for them. This becomes especially relevant in light of the development of low-cost blood tests [72, 73], advances in risk-reducing lifestyle programs [3, 74-77] and progress on disease-modifying therapies [78-81]. Our systematic review provides an in-depth overview of considerations regarding a diagnosis of AD before dementia. Strengths are our broad query to include publications from various disciplines, strict adherence to PRISMA guidelines, and use of state-of-the art methodology to inductively analyze the literature. Among the potential limitations is the restriction to articles presenting theoretical data. A next step is an inventory of empirical evidence in the clinical, personal, and societal contexts to compare expectations to experiences and identify gaps in knowledge. Immediate requirements include devising protocols for clinical practice, supportive services for patients, and legislation to protect their rights [82]. The identified considerations offer helpful starting points to prepare for a future with precision medicine and prevention of AD.

## 6 CONCLUSIONS

Diagnosing AD in individuals without dementia involves diverse and often opposing considerations. With the recent conditional approval of a first disease-modifying therapy by the FDA, there is an urgent need for empirical evidence and clinical recommendations to support practitioners, patients and society with respect to diagnosing AD before dementia.

## Data Availability

Data are available upon reasonable request.

## ACKNOWLEDGEMENTS

Research of Alzheimer center Amsterdam is part of the neurodegeneration research program of Amsterdam Neuroscience. Alzheimer Center Amsterdam is supported by Stichting Alzheimer Nederland and Stichting VUmc fonds. JvdS is appointed at ABOARD, which is a public-private partnership receiving funding from ZonMW (#73305095007) and Health∼Holland, Topsector Life Sciences & Health (PPP-allowance; #LSHM20106). More than <u>30 partners</u> participate in ABOARD. ABOARD also receives funding from Edwin Bouw Fonds and Gieskes-Strijbisfonds. PS is recipient of JPND-funded EURO-FINGERS (ZonMW-Memorabel #733051102).

